# Validation of a Multi-ancestry Polygenic Risk Score and Age-Specific Risks of Prostate Cancer: A Meta-analysis Within Diverse Populations

**DOI:** 10.1101/2022.05.03.22274606

**Authors:** Fei Chen, Burcu F Darst, Ravi K Madduri, Alex A Rodriguez, Xin Sheng, Christopher T Rentsch, Caroline Andrews, Wei Tang, Adam S Kibel, Anna Plym, Kelly Cho, Mohamed Jalloh, Serigne Magueye Gueye, Lamine Niang, Olufemi Ogunbiyi, Olufemi Popoola, Akindele O. Adebiyi, Oseremen I. Aisuodionoe-Shadrach, Hafees O Ajibola, Mustapha A Jamda, Olabode P Oluwole, Maxwell Nwegbu, Ben Adusei, Sunny Mante, Afua Darkwa-Abrahams, James E. Mensah, Andrew Anthony Adjei, Halimatou Diop, Joseph Lachance, Timothy R Rebbeck, Stefan Ambs, J. Michael Gaziano, Amy C Justice, David V Conti, Christopher A Haiman, the VA Million Veteran Program

## Abstract

**Background:** We recently developed a multi-ancestry polygenic risk score (PRS) that effectively stratifies prostate cancer risk across populations. In this study, we validated the performance of the PRS in the multi-ancestry Million Veteran Program (MVP) and additional independent studies.

**Methods:** Within each ancestry population, the association of PRS with prostate cancer risk was evaluated separately in each case-control study and then combined in a fixed-effects inverse-variance-weighted meta-analysis. We further assessed the effect modification by age and estimated the age-specific absolute risk of prostate cancer for each ancestry population.

**Results:** The PRS was evaluated in 31,925 cases and 490,507 controls, including men from European (22,049 cases, 414,249 controls), African (8,794 cases, 55,657 controls), and Hispanic (1,082 cases, 20,601 controls) populations. Comparing men in the top decile (90-100% of the PRS) to the average 40-60% PRS category, the prostate cancer OR was increased 3.8-fold in European ancestry men (95% CI=3.62-3.96), 2.8-fold in African ancestry men (95% CI=2.59-3.03), and 3.2-fold in Hispanic men (95% CI=2.64-3.92). The PRS did not discriminate risk of aggressive versus non-aggressive prostate cancer. However, the OR diminished with advancing age (European ancestry men in the top decile: ≤55 years, OR=7.11; 55-60 years, OR=4.26; >70 years, OR=2.79). Men in the top PRS decile reached 5% absolute prostate cancer risk ∼10 years younger than men in the 40-60% PRS category.

**Conclusions:** Our findings validate the multi-ancestry PRS as an effective prostate cancer risk stratification tool across populations. A clinical study of PRS is warranted to determine if the PRS could be used for risk-stratified screening and early detection.

**Funding:** This work was supported by the National Cancer Institute at the National Institutes of Health (grant numbers U19 CA214253 to C.A.H., U01 CA257328 to C.A.H., U19 CA148537 to C.A.H., R01 CA165862 to C.A.H., K99 CA246063 to B.F.D, and T32CA229110 to F.C), the Prostate Cancer Foundation (grants 21YOUN11 to B.F.D. and 20CHAS03 to C.A.H.), the Achievement Rewards for College Scientists Foundation Los Angeles Founder Chapter to B.F.D, and the Million Veteran Program-MVP017. This research has been conducted using the UK Biobank Resource under application number 42195. This research is based on data from the Million Veteran Program, Office of Research and Development, and the Veterans Health Administration. This publication does not represent the views of the Department of Veteran Affairs or the United States Government.

## Introduction

Prostate cancer is the second leading cause of cancer death and represents one of the largest health disparities in the US, with African ancestry men having the highest incidence rates(1). Genetic factors play an important role in prostate cancer susceptibility(2, 3) and racial/ethnic disparities in disease incidence(3). We recently developed a multi-ancestry polygenic risk score (PRS) that effectively stratifies prostate cancer risk across populations(3). The PRS could potentially be an effective tool to identify men across diverse populations at higher risk of developing prostate cancer and allow them to make more informed decisions regarding at what age(s) and how frequently to undergo PSA screening.

In this investigation, we evaluated the previously developed multi-ancestry PRS in large independent samples of men from the Veteran Affairs Million Veteran Program (MVP; 21,078 cases and 284,177 controls, including 13,643 cases and 210,214 controls of European ancestry, 6,353 cases and 53,362 controls of African ancestry, and 1,082 cases and 20,601 controls from Hispanic populations)(4), the Men of African Descent and Carcinoma of the Prostate (MADCaP) Network (405 cases and 396 controls of African ancestry)(5), and the Maryland Prostate Cancer Case-Control Study (NCI-MD; 383 cases and 395 controls of African ancestry)(6) (**Materials and Methods**). We also included, through meta-analysis, independent replication studies of the multi-ancestry PRS conducted to date in European (UK Biobank and Mass General Brigham [MGB] Biobank) and African ancestry populations (California and Uganda Prostate Cancer Study [CA UG] and MGB Biobank; **Materials and Methods**)(3, 7), bringing the total sample to 31,925 cases and 490,507 controls.

## Results

The multi-ancestry PRS was strongly associated with prostate cancer risk in the three populations (**Figure 1** and **Figure 1 – source data 1**). In European ancestry men, ORs were 3.78 (95% CI=3.41-3.81) and 7.32 (95% CI=6.76-7.92) for men in the top PRS decile (90-100%) and top percentile (99-100%), respectively, compared to men with average genetic risk (40-60% PRS category). In African ancestry men, ORs were 2.80 (95% CI=2.49-2.95) and 4.98 (95% CI=4.27-5.79) for men in the top PRS decile and percentile, respectively. In Hispanic men, ORs were 3.22 (95% CI=2.64-3.92) and 6.91 (95%=4.97-9.60) for men in the top PRS decile and percentile, respectively. PRS associations within populations were generally consistent across individual replication studies (**Figure 1 – figure supplement 1**). The area under the curve (AUC) increased 0.136 on average across populations upon adding the PRS to a base model of age and principal components of ancestry (**Appendix 1 - Table 1**). Compared to the mean PRS in European ancestry controls, African ancestry controls had a mean PRS associated with a relative risk of 2.19 (95% CI=2.17-2.21), while Hispanic controls had a relative risk of 1.16 (95% CI=1.15-1.18), consistent with previous findings(3).

**Figure 1.**
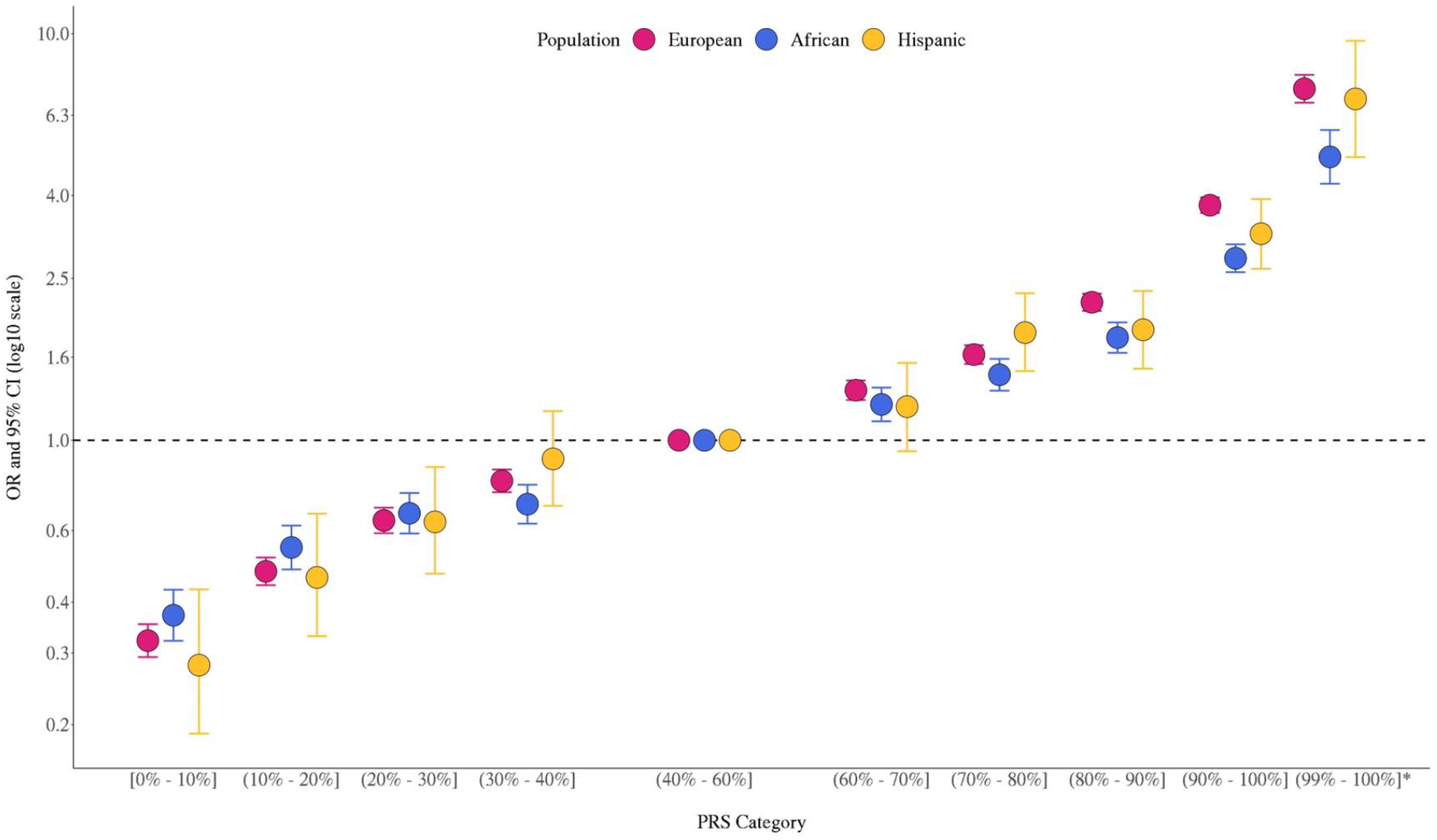
Association between the multi-ancestry PRS of 269 variants and prostate cancer risk in men from European, African, and Hispanic populations. The European ancestry replication studies included MVP, the UK Biobank (Conti, Darst et al., *Nature Genetics*, 2021), and the Mass General Brigham Biobank (MGB; Plym et al., *JNCI*, 2021). The African ancestry replication studies included MVP, the California and Uganda Prostate Cancer Study (CA UG; Conti, Darst et al., *Nature Genetics*, 2021), the Men of African Descent and Carcinoma of the Prostate (MADCaP) Network, the Maryland Prostate Cancer Case-Control Study (NCI-MD), and MGB (Plym et al., *JNCI*, 2021). Replication in Hispanic men was conducted in MVP. Results from individual replication studies are shown in **Figure 1 – figure supplement 1**. The x-axis indicates the PRS category. Additional analysis was performed to evaluate the PRS association in men with extremely high genetic risk (99%-100%). The y-axis indicates odds ratio (OR) with error bars representing 95% confidence interval (CI) for each PRS category compared to the 40-60% PRS. The dotted horizontal line corresponds to an OR of 1. ORs and 95% CIs for each decile are provided in **Figure 1 – source data 1**.

Previously, we found that PRS associations were significantly stronger in younger men (aged ≤55 years) than in older men (aged >55 years)(3). In the two large replication studies, UK Biobank and MVP, we further explored effect modification by age (**Figure 2, Figure 2 – figure supplement 1**, and **Figure 2 – source data 1**). In European ancestry men, for the top PRS decile, the OR was 7.11 (95% CI=5.82-8.70) in men aged ≤55, 4.26 (95% CI=3.77-4.81) in men aged 55-60, and 2.79 (95% CI=2.50-3.11) in men aged >70. The gradient in PRS risk by age was greater for men in the top PRS percentile, with ORs of 17.2 (95% CI=13.0-22.8), 9.18 (95% CI=7.52-11.2), and 5.43 (95% CI=4.50-6.55) estimated for men ≤55, 55-60, and >70 years of age, respectively. Attenuation of PRS associations with age was also observed in African ancestry men, as the OR for men in the top PRS decile decreased from 3.75 (95% CI=3.04-4.64) in men aged ≤55 to 2.16 (95% CI=1.76-4.68) in men aged >70. For African ancestry men in the top PRS percentile, the OR decreased from 8.80 (95% CI=6.16-12.6) in men aged ≤55 to 2.87 (95% CI=1.76-4.68) in men aged >70. A similar trend was observed in Hispanic men (OR=6.37, 95% CI=3.26-12.44 for men ≤55 and OR=2.15, 95% CI=1.39-3.32 for men >70 in the top PRS decile). Compared to men in the 40-60% PRS category, men from European, African, and Hispanic populations in the top PRS decile reached 5% absolute risk of prostate cancer 12 years earlier (age 57 versus 69), 8 years earlier (age 55 versus 63), and 11 years earlier (age 60 versus 71), respectively (**Table 1** and **Figure 3**). For men in the top PRS percentile, 5% absolute risk was reached by ages 51, 52, and 53 for European, African, and Hispanic populations, respectively.

**Figure 2.**
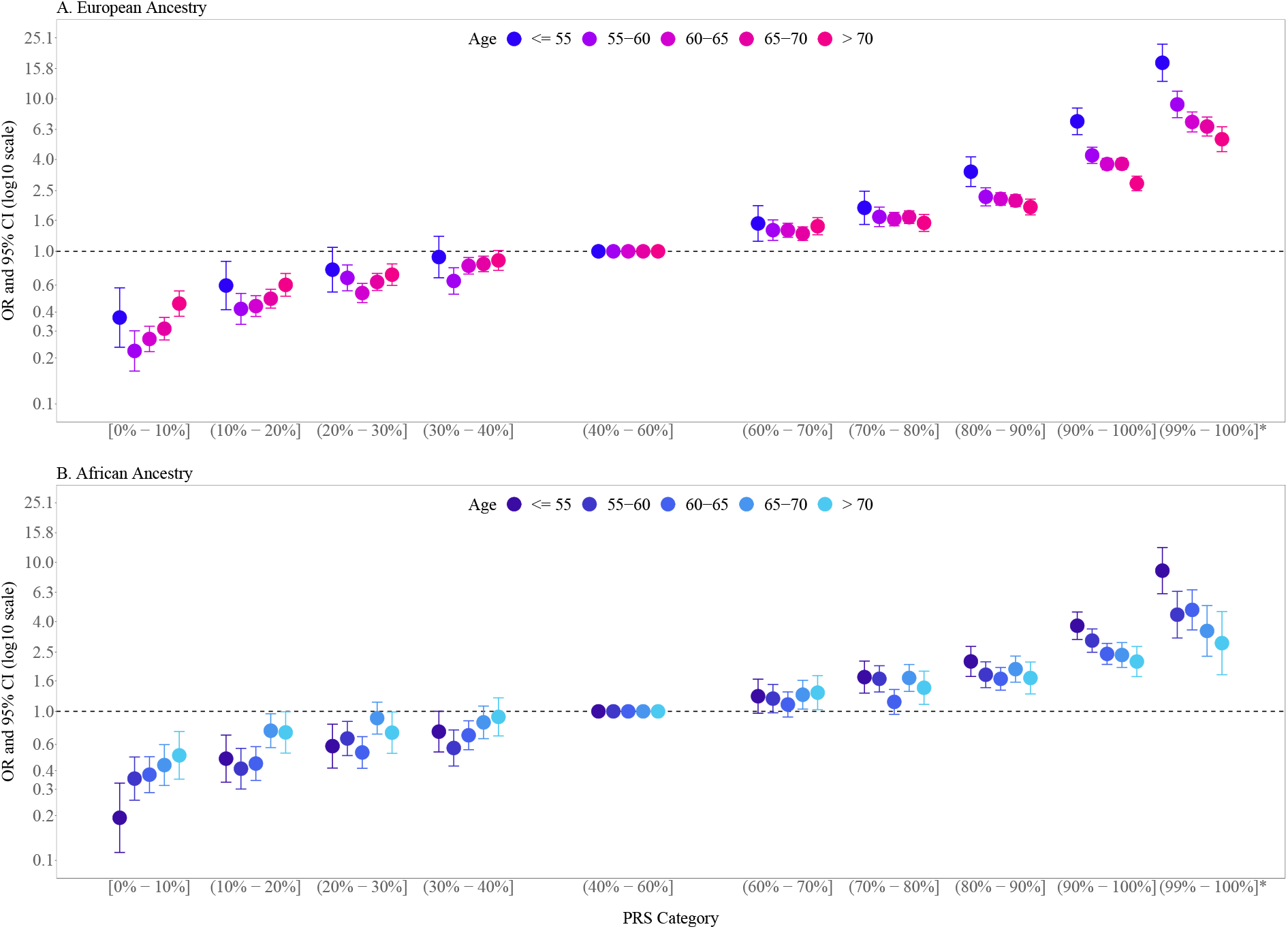
Association between the multi-ancestry PRS of 269 variants and prostate cancer risk stratified by age. PRS associations in men of European ancestry (A) were meta-analyzed from UK Biobank (6,852 cases and 193,117 controls) and MVP (13,643 cases and 210,214 controls; **Figure 2 – figure supplement 1**), whereas PRS associations in men of African ancestry (B) were estimated from MVP (6,353 cases and 53,362 controls). The x-axis indicates the PRS category. Additional analyses were performed to evaluate the PRS association in men with extremely high genetic risk (top percentile, 99%-100%). The y-axis indicates the odds ratio (OR) with error bars representing the 95% confidence interval (CI) for each PRS category compared to the 40-60% PRS category. The dotted horizontal line corresponds to an OR of 1. ORs and 95% CIs for each PRS category are provided in **Figure 2 – source data 1**.

**Table 1.**
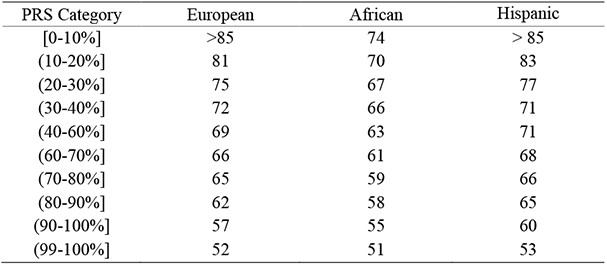
Age at which 5% absolute risk of prostate cancer is reached in men from European, African, and Hispanic populations. Absolute risks of prostate cancer were estimated using age- and population-specific Surveillance, Epidemiology, and End Results (SEER) incidence rates, CDC National Center for Health Statistics mortality rates, and PRS associations from **Supplementary File 2 - Table S1** based on MVP and the UK Biobank.

**Figure 3.**
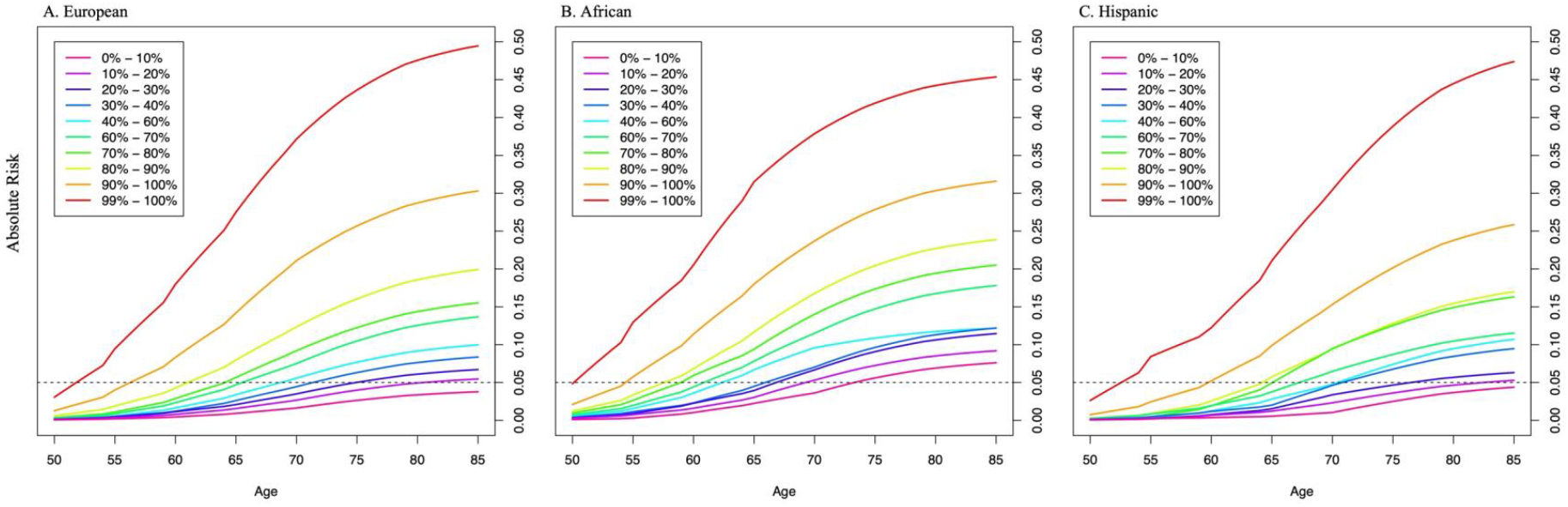
Absolute risk of prostate cancer by PRS category in men from European (A), African (B), and Hispanic populations (C). The absolute risks were estimated using the age- and population-specific PRS associations from **Figure 2 – source data 1**, the SEER incidence rates, and the CDC mortality rates corresponding to non-Hispanic White, Black, and Hispanic men. The dotted line indicates the 5% absolute risk of prostate cancer.

Similar to previous findings(3, 7), the multi-ancestry PRS did not consistently differentiate aggressive and non-aggressive prostate cancer risk (**Appendix 1 - Table 2**). For men in the top PRS decile, ORs were 3.17 (95% CI=2.77-3.63) and 3.71 (95% CI=3.48-3.94) for aggressive and non-aggressive prostate cancer, respectively, in European ancestry men (P-heterogeneity=0.04) and 1.92 (95% CI=1.17-3.15) and 3.30 (95% CI=2.64-4.12), respectively, in Hispanic men (P-heterogeneity=0.05). In African ancestry men, the association was greater for aggressive (OR=3.31, 95% CI=2.71-4.03) than non-aggressive disease (OR=2.66, 95% CI=2.43-2.92), although confidence intervals overlapped (P-heterogeneity=0.05).

## Discussion

Findings from this investigation provide further support for the PRS as a prostate cancer risk stratification tool in men from European, African, and Hispanic populations. Notably, this investigation provides the first evidence of replication of the multi-ancestry PRS in Hispanic men. Consistent with previous findings(3, 7), we observed lower PRS performance in African versus European ancestry men, supporting the need to expand GWAS and fine-mapping efforts in African ancestry men. The stronger association of the PRS with prostate cancer risk observed for younger men supports previous studies(3), suggesting that the contribution of genetic factors to prostate cancer is greater at younger ages and that age needs to be considered when comparing PRS findings across studies and populations.

The PRS is an effective risk stratification tool for prostate cancer at both ends of the risk spectrum. Current guidelines consider age, self-reported race, and a family history of prostate cancer in PSA screening decisions(8). Although the PRS generally did not differentiate aggressive versus non-aggressive prostate cancer, a substantial fraction of men who will develop aggressive tumors (∼40%) are among a subset of men in the population with the highest PRS (top 20%), while only ∼7% of men who will develop aggressive tumors are among the subset of men in the population with the lowest PRS (bottom 20%), suggesting that reduced screening among low PRS men may reduce the overdiagnosis of prostate cancer. Indeed, previous studies in men of European ancestry support that PRS-stratified screening could significantly reduce the overdiagnosis of prostate cancer by 33%-42%, with the largest reduction observed in men with lower genetic risk(9-11). Risk-stratified screening studies are warranted in diverse populations to evaluate the clinical utility of the multi-ancestry PRS for early disease detection and when in a man’s life genetic risk should be considered in the shared decision-making process for prostate cancer screening.

## Materials and Methods

### Participants and Genetic Data

We replicated the association between the multi-ancestry PRS and prostate cancer risk in three independent case-control samples from the VA Million Veteran Program (MVP), the Men of African Descent and Carcinoma of the Prostate (MADCaP) Network, and the Maryland Prostate Cancer Case-Control Study (NCI-MD), as described below. Previously, this multi-ancestry PRS was replicated by our group and others in the California and Uganda Prostate Cancer Study (CA UG, 1,586 cases and 1,047 controls of African ancestry), the UK Biobank (6,852 cases and 193,117 controls of European ancestry; updates to the UK Biobank led to slightly different sample sizes in the present study of 8,483 cases and 193,744 controls of European ancestry), and the Mass General Brigham Biobank (MGB, formerly known as the Partners Healthcare Biobank, 67 cases and 457 controls of African ancestry and 1,554 cases and 10,918 controls of European ancestry). Results from these studies are described in detail elsewhere(3, 7). To provide a comprehensive assessment of the PRS validation, we meta-analyzed all replication studies, which included a total of 22,049 cases and 414,249 controls of European ancestry (UK Biobank, MGB, and MVP) and 8,794 cases and 55,657 controls of African ancestry (MGB, MADCaP, NCI-MD, and MVP). In men of Hispanic ancestry, the multi-ancestry PRS was only assessed in MVP (1,082 cases and 20,601 controls). All study protocols were approved by each site’s Institutional Review Board in accordance with the principles outlined in the Declaration of Helsinki.

#### MVP

The design of the MVP has been previously described(4). Briefly, participants were recruited from approximately 60 Veteran Health Administration (VHA) facilities across the United States since 2011 with the current enrollment at >800,000. Informed consent is obtained for all participants to provide a blood sample for genetic analysis and to access their full clinical and health data. A total of 485,856 samples from participants enrolled between 2011 and 2017 were genotyped on a custom Axiom array designed specifically for MVP (MVP 1.0). The genotyping array design and data quality controls were extensively described elsewhere(12). After excluding variants with high genotype missingness (>5%) and those deviated from the expected allele frequency observed in the reference populations, genotype data was imputed to the 1000 Genomes Project Phase 3 reference panel(13). In MVP, genetic ancestry was assessed using HARE(14), which assigned >98% of participants with genotype data to one of four non-overlapping population groups: non-Hispanic White (European), non-Hispanic Black (African), Hispanic, and non-Hispanic Asian. Due to the small number of non-Hispanic Asian individuals, they are excluded from the current analysis.

We identified a total of 21,078 cases and 284,177 controls from MVP, of whom 13,643 cases and 210,214 controls were of European ancestry (73.3%), 6,353 cases and 53,362 controls were of African ancestry (19.6%), and 1,082 cases and 20,601 controls were Hispanic (7.1%). Prostate cancer cases were identified from the Veterans Affairs Central Cancer Registry (VACCR), which collects cancer diagnosis, extent of disease and staging, first course of treatment, and outcomes from 132 VA medical centers. In this analysis, we only included cases from the VACCR who have a confirmed cancer diagnosis based on their diagnostic code, procedure code, and information from other clinical documents. Among the MVP participants without any prostate cancer diagnostic codes, we limited controls to those aged 45 to 95 years and had at least one prostate-specific antigen (PSA) test after enrollment. For prostate cancer cases, we obtained additional information on cancer staging and Gleason score to define aggressive prostate cancer phenotypes. Specifically, we defined aggressive prostate cancer as tumor stage T3/T4, regional lymph node involvement (N1), metastatic disease (M1), or Gleason score ≥8.0, while non-aggressive cases were defined as tumor stage T1/T2 and Gleason score <7.

#### MADCaP

The MADCaP Network dataset included 405 prostate cancer cases and 396 controls from sub-Saharan Africa, as previously described(5, 15), with a substantial proportion of cases diagnosed at late stages. The MADCaP samples were genotyped on a customized array designed to capture common genetic variation in diverse African populations, and genotyping and quality control have been described in detail elsewhere(5). GWAS data were imputed using the 1000 Genomes Project Phase 3 reference panel(13).

#### NCI-MD

The NCI-MD Study included 383 prostate cancer cases identified from two Maryland hospitals and 395 population-based controls from Maryland and its neighboring states(16). About 87% of the cases in this study were considered non-aggressive, with pathologically confirmed T1 or T2 tumor and a Gleason score ≤7. All samples from this study were genotyped on the Illumina InfiniumOmni5Exome array and were imputed to the 1000 Genomes Project Phase 3 reference panel(13).

### PRS Construction and Association Analyses

PRSs were constructed by summing variant-specific weighted allelic dosages from 269 previously identified prostate cancer risk variants(3). Variants were weighted using the multi-ancestry conditional weights generated from our previous trans-ancestry genome-wide association study (GWAS) for prostate cancer (3). Variants and weights used to generate the PRS can be found in the PGS Catalog: https://www.pgscatalog.org/publication/PGP000122/.

The association of PRS on prostate cancer risk was estimated separately in each ancestry population using an indicator variable for the percentile categories of the PRS distribution: [0-10%], (10-20%], (20-30%], (30-40%], (40-60%], (60-70%], (70-80%], (80-90%], and (90-100%], where parentheses indicate greater than and square brackets indicate less than or equal to. Additional analysis was performed to obtain the association for the top 1% PRS by splitting the top PRS decile into (90%-99%] and (99%-100%] categories. PRS thresholds were determined in the observed distribution among controls. In all replication studies, logistic regression was performed to estimate ORs corresponding to each PRS category, adjusting for age and the up to ten principal components of ancestry, with the (40-60%] category as the reference. Age was defined as age at diagnosis for prostate cancer cases and age at last PSA testing (MVP) or age at study recruitment (MADCaP and NCI-MD) for controls.

Discriminative ability was evaluated in MVP by estimating the area under the curve (AUC) for logistic regression models of prostate cancer that included covariates only (age and four principal components of ancestry) and for models that additionally included the PRS. All analyses were performed separately within each population.

We performed a fixed-effects inverse-variance-weighted meta-analysis to combine the ORs and standard errors for each PRS decile from individual replication studies by ancestry using R package *meta*. This meta-analysis was conducted across the three studies of European ancestry, UK Biobank, MGB, and MVP, as well as across the four studies of African ancestry, MGB, MADCaP, NCI-MD, and MVP.

In the two large replication studies, UK Biobank and MVP, logistic regression analyses were repeated stratifying both cases and controls at ages ≤55, (55-60], (60-65], (65-70], and >70, with adjustments for age (as a continuous variable) and the top principal components of ancestry.

The PRS associations estimated in men of European ancestry from UK Biobank and MVP were meta-analyzed using a fixed-effects inverse-variance-weighted method. Heterogeneity between studies and across strata was assessed via a Q statistic between effects estimates with corresponding tests of significance.

In the three ancestry populations from MVP, we also performed stratified analyses by disease aggressiveness, where cases were stratified as aggressive or non-aggressive and all controls were used in corresponding stratified analysis. Heterogeneity across strata was assessed via a Q statistic between effects estimates with corresponding tests of significance.

### Estimation of Absolute Risk

The absolute risk of prostate cancer was calculated for a given age for each PRS category in European, African, and Hispanic ancestry men(17-20). The approach constrains the PRS-specific absolute risks for a given age to be equivalent to the age-specific incidences for the entire population, such that age-specific incidence rates are calculated to increase or decrease based on the estimated risk of PRS category and the proportion of the population within the PRS category. The calculation accounts for competing causes of death.

Specifically, for a given population and PRS category *k* (e.g., 80-90%, 90-100%), the absolute risk by age *t* is computed as: 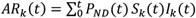. This calculation consists of three components:

1. ***P***_*ND*_(*t*) is the probability of not dying from another cause of death by age *t* using age-specific mortality rates, 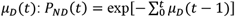. Age-specific mortality rates are provided from a reference cohort.
2. *S*_*k*_(*t*) is the probability of surviving prostate cancer by age *t* in the PRS category *k* and uses the prostate cancer incidence by age *t* for category 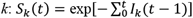.
3. The prostate cancer incidence by age *t* for PRS category *k* is *I*_*k*_(*t*) and is calculated by multiplying the population prostate cancer incidence for the reference category, *I*_0_(*t*) and the corresponding risk ratio, *β*_*ka*_, for PRS category *k* and age category *a* (e.g. ages ≤55, 55-60, 60-65, 65-70, and >70) containing age *t*. These are estimated from the odds ratio obtained from the population-specific individual-level PRS analysis for each age-strata (African and Hispanic ancestry odds ratios from MVP and European ancestry odds ratios meta-analyzed from MVP and UK Biobank): *I*_*k*_(*t*) = *I*_0_(*t*)exp (*β*_*ka*_).

Prostate cancer incidence for age *t* for the reference category, *I*_0_(*t*)’ is obtained by constraining the weighted average of the population cancer incidences for the PRS categories to the population age-specific prostate cancer incidence, 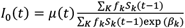, where *f*_*k*_ is the frequency of the PRS category *k* with *f*_*k*_ = 0.1 for all non-reference categories in our primary PRS analysis by deciles (e.g., 0-10%, 10-20%, 20-30%, etc.).

By leveraging the definition that *S*_*k*_(*t* = 0) = 1, for all *k*, the absolute risks were calculated iteratively by first getting *I*_0_(*t* = 1), then *I*_*k*_(*t* = 1), then *S*_*k*_(*t* = 1) and finally *AR*_*k*_ (*t* = 1). Subsequent values were then calculated recursively for all *t*.

For each population, absolute risks by age *t* were calculated using age- and population-specific prostate cancer incidence, *μ*(*t*), from the Surveillance, Epidemiology, and End Results (SEER) Program (1999-2013) and age- and population-specific mortality rates, *μ*_*D*_(*t*), from the National Center for Health Statistics, CDC (1999-2013).

## Data Availability

This investigation included published results from the following studies under DOI numbers 10.1038/s41588-020-00748-0 and 10.1093/jnci/djab058. The MVP individual level data is available to approved VA researchers through standard mechanisms. Full GWAS summary statistics can be found in dbGaP (https://www.ncbi.nlm.nih.gov/gap/) under the MVP accession (phs001672).Publicly available data described in this manuscript can be found from the following websites: 1000 Genomes Project (https://www.internationalgenome.org/); SEER (https://seer.cancer.gov/); National Center for Health Statistics, and CDC (https://www.cdc.gov/nchs/index.htm).

## Data Availability Statement

### Data availability

This investigation included published results from the following studies under DOI numbers <u>10.1038/s41588-020-00748-0</u> and <u>10.1093/jnci/djab058</u>. The MVP individual level data is available to approved VA researchers through standard mechanisms. Full GWAS summary statistics can be found in dbGaP (https://www.ncbi.nlm.nih.gov/gap/) under the MVP accession (phs001672).Publicly available data described in this manuscript can be found from the following websites: 1000 Genomes Project (https://www.internationalgenome.org/); SEER (https://seer.cancer.gov/); National Center for Health Statistics, and CDC (https://www.cdc.gov/nchs/index.htm).

### Code availability

All analyses were performed using R statistical packages freely available at https://cran.r-project.org/mirrors.html. The R code for the PRS association analysis was modified from the code available at https://github.com/USCmec/Polfus_Darst_HGGA_2021/. Source data for Figure 1 and Figure 2 are provided.

## FIGURES AND TABLES

**Figure 1 – figure supplement 1.**
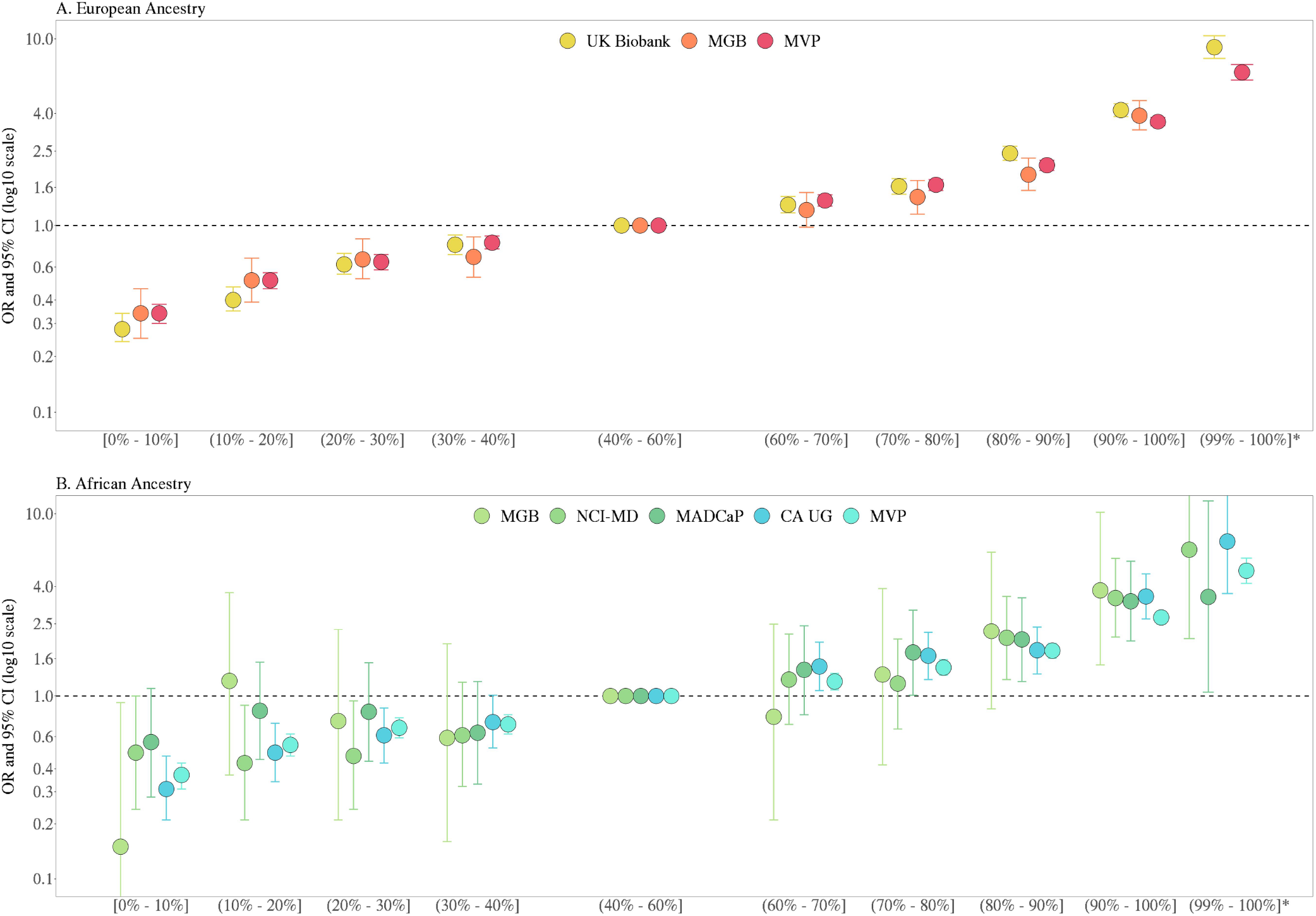
Association between the multi-ancestry PRS of 269 variants and prostate cancer risk from individual replication studies of European (A) and African ancestry (B). Replication studies in men of European and African ancestry included MVP (13,643 cases and 210,214 controls of European ancestry, 6,353 cases and 53,362 controls of African ancestry), the UK Biobank (6,852 cases and 193,117 controls of European ancestry), the Mass General Brigham Biobank (MGB; 67 cases and 457 controls of African ancestry and 1,554 cases and 10,918 controls of European ancestry), the California and Uganda Prostate Cancer Study (CA UG; 1,586 cases and 1,047 controls of African ancestry), the Men of African Descent and Carcinoma of the Prostate (MADCaP) Network (405 cases and 396 controls of African ancestry), and the Maryland Prostate Cancer Case-Control Study (NCI-MD; 383 cases and 395 controls of African ancestry). The x-axis indicates the PRS category. Additional analysis was performed to evaluate the PRS association in men with extremely high genetic risk (99%-100%) in all individual studies except MGB. The y-axis indicates odds ratio (OR) with error bars representing the 95% confidence interval (CI) for each PRS category compared to the 40-60% PRS category. The dotted horizontal line corresponds to an OR of 1.

**Figure 1 – source data 1**

**Association between the multi-ancestry PRS and prostate cancer risk replicated in men from European, African, and Hispanic populations**. Results in men of European ancestry were meta-analyzed across MVP, UK Biobank, and Partners. Results in men of African ancestry were meta-analyzed across MVP, CA UG, NCI-MD, and MADCaP. Results in Hispanic men were from MVP. The PRS association for men in the 99-100% category was not assessed in Partners and therefore was not included in the meta-analysis. PRS categories were determined based on the distribution in controls. Odds ratio (OR) and 95% confidence interval (95% CI) were estimated from logistic regression models adjusting for age and principal components of ancestry.

**Figure 2 – figure supplement 1.**
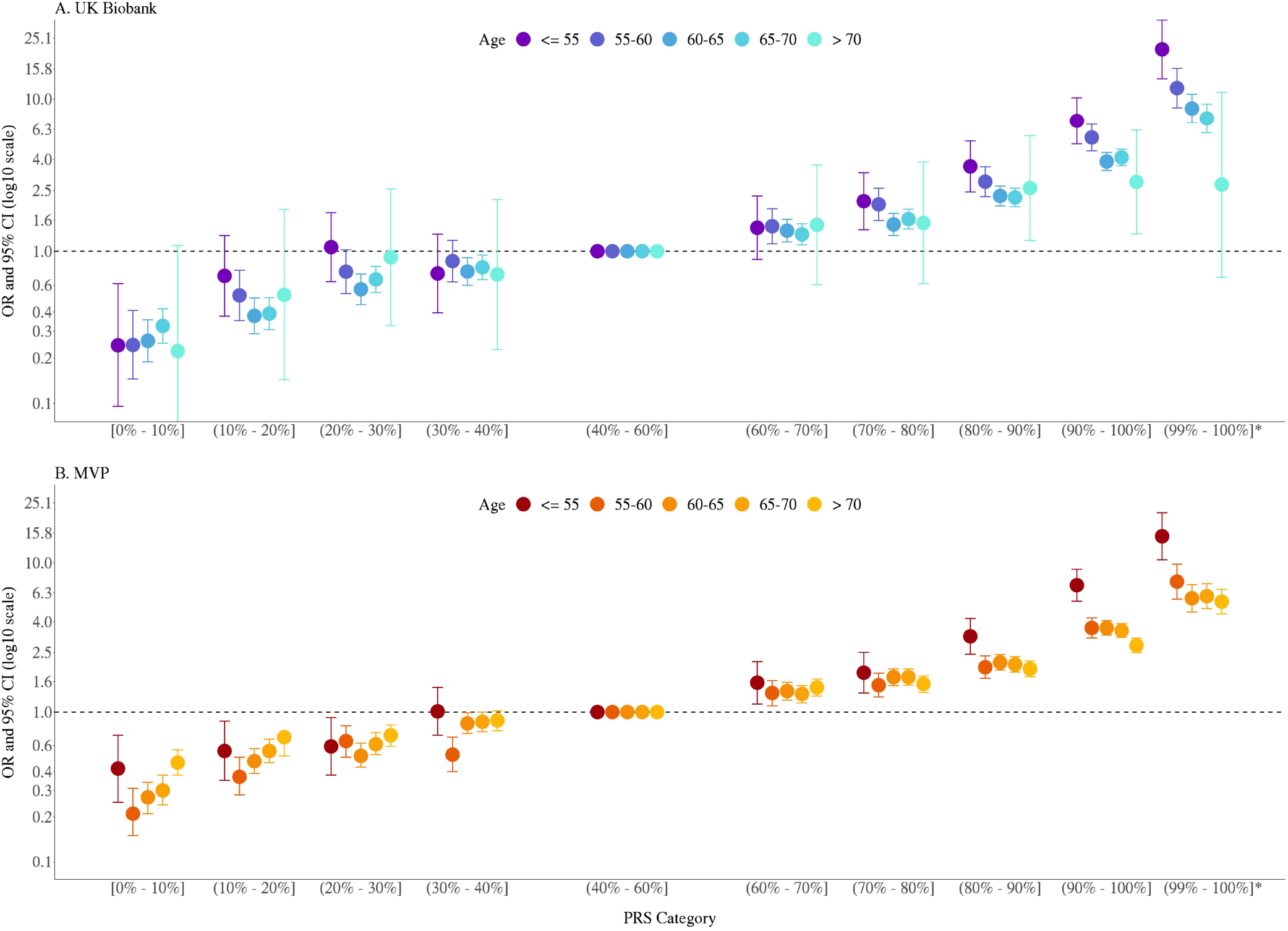
Association between the multi-ancestry PRS of 269 variants and prostate cancer risk stratified by age in men of European ancestry from UK Biobank (A) and MVP (B). The x-axis indicates the PRS category. Additional analysis was performed to evaluate the PRS association in men with extremely high genetic risk (99%-100%). The y-axis indicates odds ratio (OR) with error bars representing the 95% confidence interval (CI) for each PRS category compared to the 40-60% PRS category. The dotted horizontal line corresponds to an OR of 1.

**Figure 2 – source data 1**

**Association of multi-ancestry PRS and prostate cancer risk stratified by age**. Results in men of European ancestry were meta-analyzed across UK Biobank and MVP while results in men of African and Hispanic ancestry were estimated in MVP only. PRS categories were determined based on the distribution in controls. Odds ratio (OR) and 95% confidence interval (95% CI) were estimated from logistic regression models adjusting for age and principal components of ancestry.

**Table 1**

**Age at which 5% absolute risk of prostate cancer is reached in men from European, African, and Hispanic populations**. Absolute risks of prostate cancer were estimated using age- and population-specific Surveillance, Epidemiology, and End Results (SEER) incidence rates, CDC National Center for Health Statistics mortality rates, and PRS associations from **Figure 2 – source data 1** based on MVP and the UK Biobank.

**Appendix 1 – Table 1.**
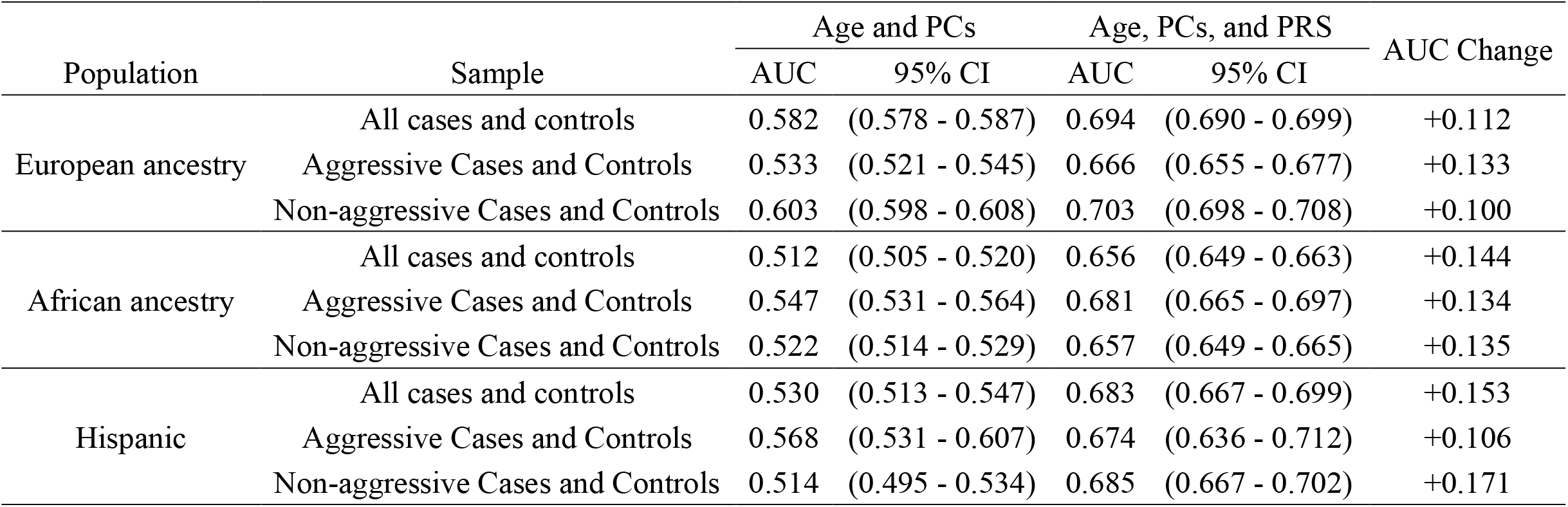
Model discrimination and improvement estimated with area under the curve (AUC) upon adding the multi-ancestry PRS to a base model in the MVP study populations.

**Appendix 1 - Table 2.**
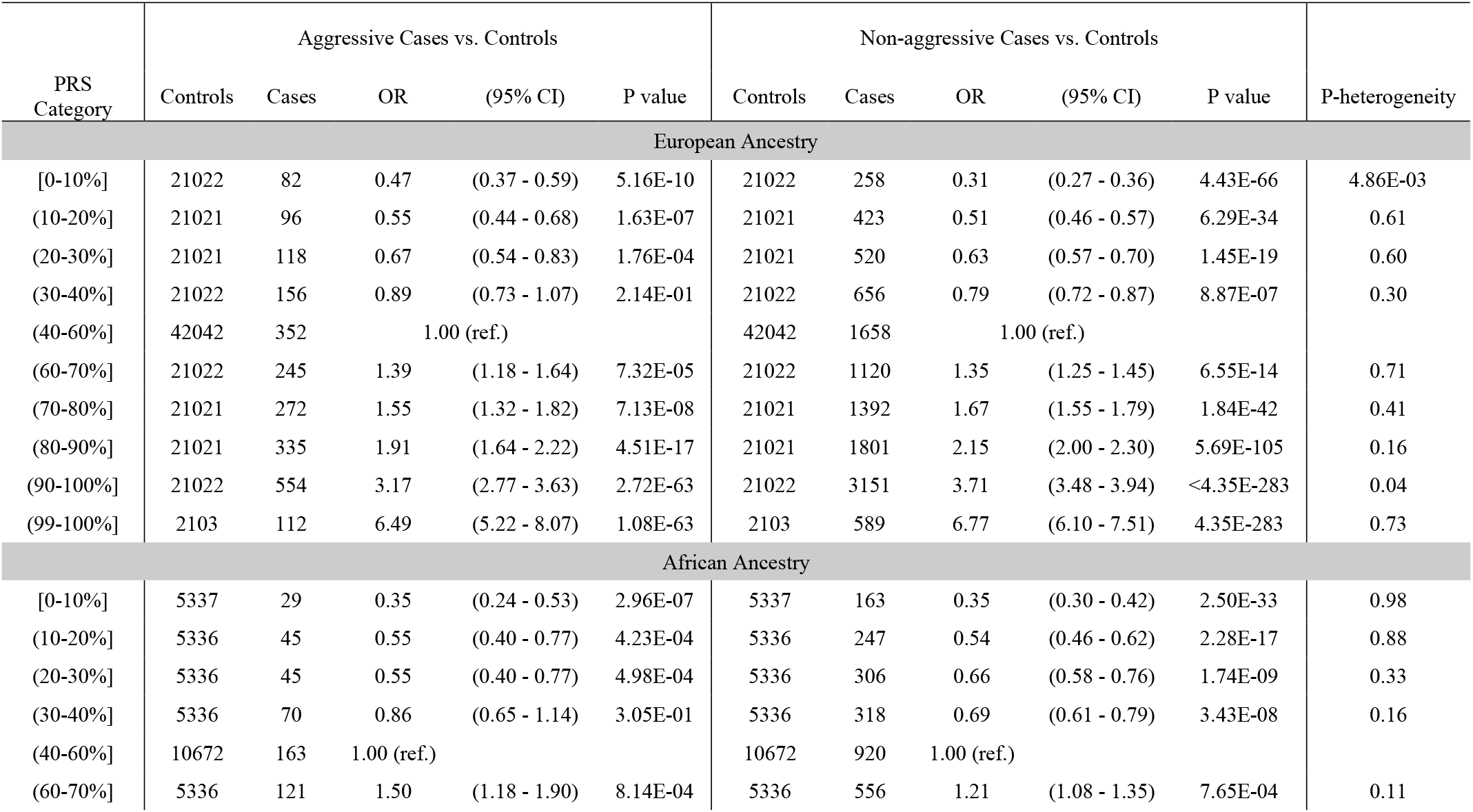
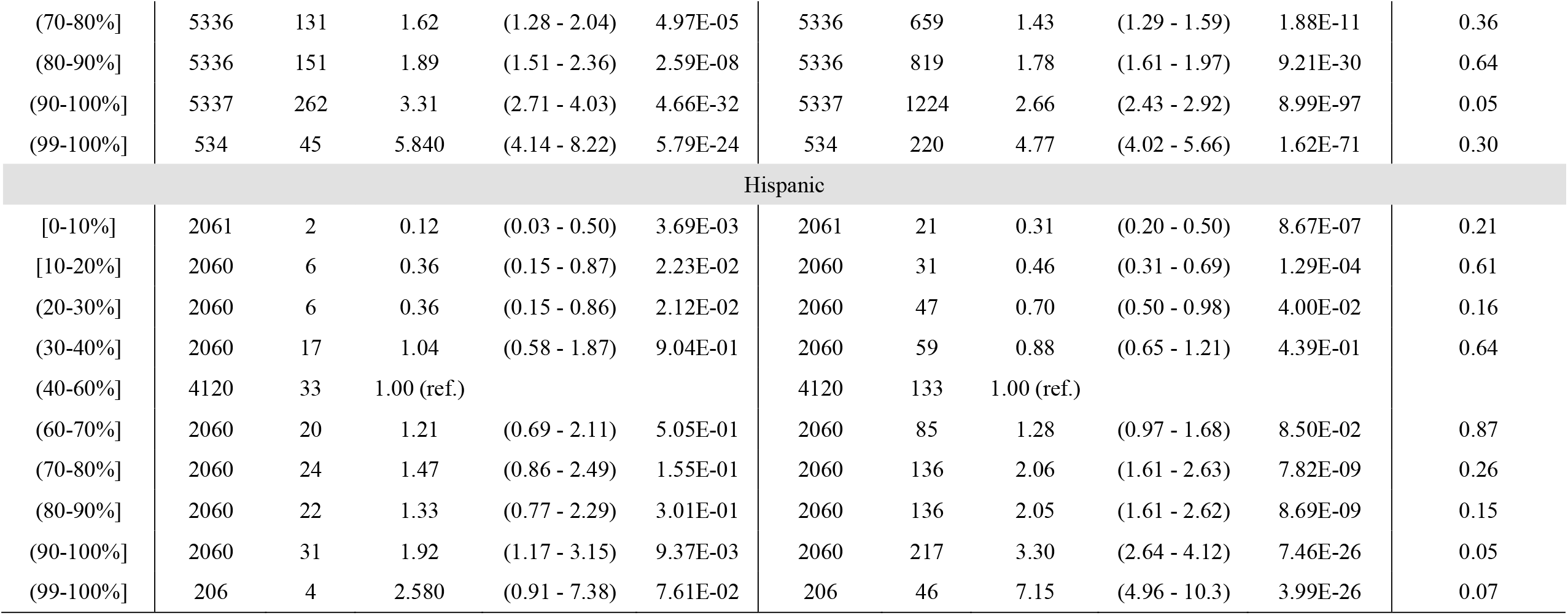
The association between the multi-ancestry PRS and prostate cancer aggressiveness in MVP participants from European, African, and Hispanic populations. PRS categories were determined based on the distribution in controls. Odds ratio (OR) and 95% confidence interval (95% CI) were estimated from logistic regression models adjusting for age and principal components of ancestry. Heterogeneity was assessed via a Q statistic between effects estimates with corresponding tests of significance.

**Appendix 1 – Figure 1.**
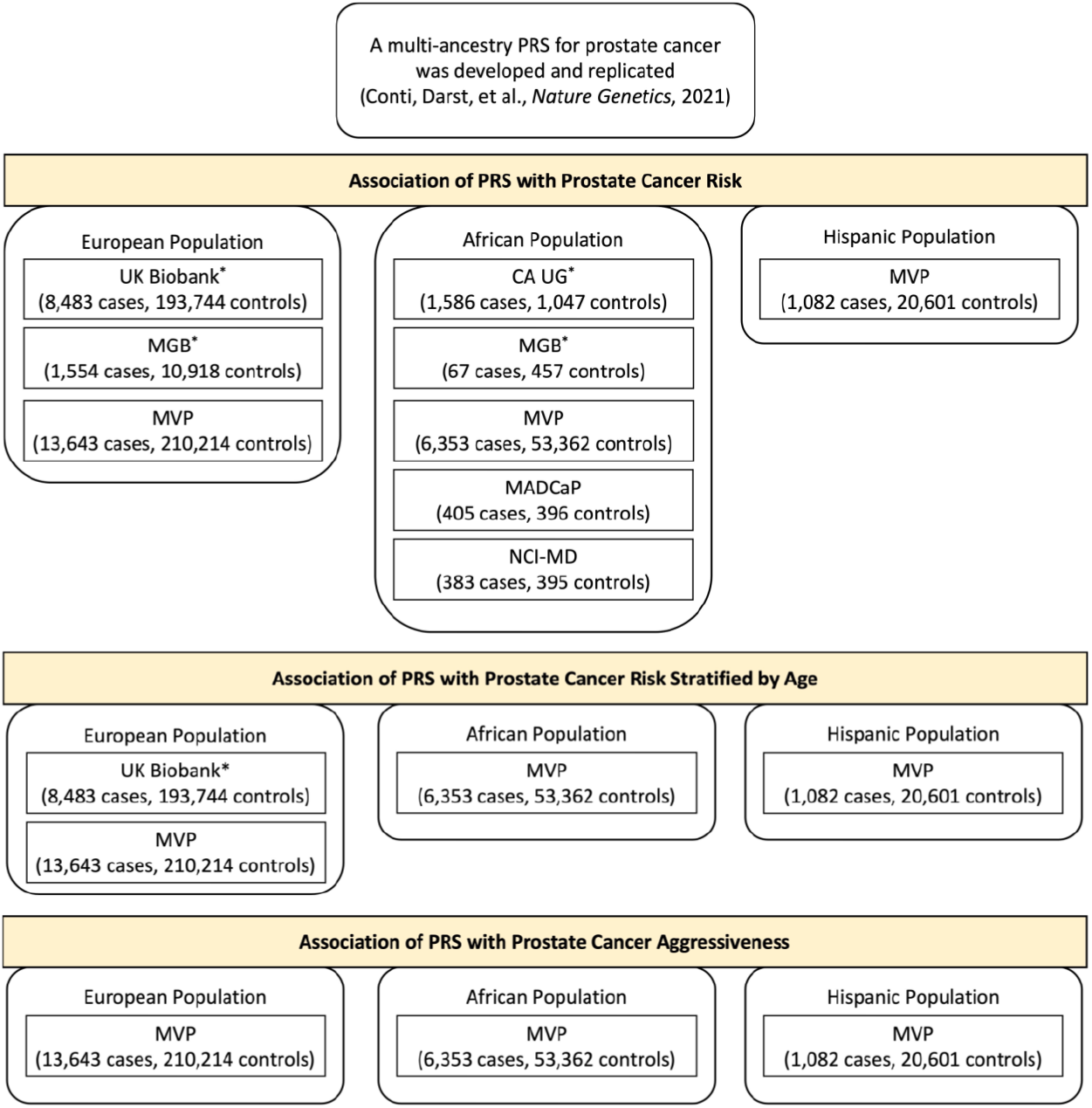
Individual studies of European, African, or Hispanic population included in the association analysis. Results from previous replication studies (*) in UK Biobank, MGB, and CA UG were meta-analyzed with results from MVP, NCI-MD and MADCaP within each population.

